# Glycated haemoglobin measurements from UK Biobank are different to those in linked primary care records: implications for combining biochemistry data from research studies and routine clinical care

**DOI:** 10.1101/2021.11.20.21266511

**Authors:** Katherine G Young, Timothy J McDonald, Beverley M Shields

**Author notes:** Corresponding Author: Level 3 RILD Building, RD&E Wonford, Barrack Road, Exeter, Devon, EX2 5DW, UK.

## Abstract

Data linkage of cohort or RCT data with routinely collected data is becoming increasingly commonplace, and this often involves combining biomarker measurements from different sources. However, sources may have different biases due to differences in assay method and calibration. Combining these measurements, or diagnoses based on these measurements, is therefore not always valid. We highlight an example using glycated haemoglobin A1c (HbA1c) test results from two different sources in UK Biobank data.

---

Biochemical tests of the same individual carried out on different test platforms are often not comparable due to bias in assay method and calibration^1, 2^. Combining measurements from different sources, or diagnoses based on these measurements, is therefore not always valid. We highlight an example using glycated haemoglobin A1c (HbA1c) test results from two different sources in UK Biobank data: HbA1c measurements taken at baseline assessment using a single assay method (the Bio-Rad Variant II Turbo HPLC analyser^3^); and HbA1c measurements from linked UK primary care records, where assay method was dependent on which NHS laboratory the sample was processed in.

We identified UK Biobank participants with no pre-existing or previous diagnosis of diabetes mellitus (any type), with a primary care HbA1c measurement ≤ 100 days before or after baseline assessment (*n*=1,039; a detailed method is provided in Supplementary Figure S1). In individuals without diabetes, HbA1c should be relatively stable within this short timeframe. We found that UK Biobank baseline measurements were on average lower than primary care measurements with a mean difference of 2 mmol/mol (Figure 1), regardless of whether the primary care measurement was taken before or after baseline assessment.

**Figure 1.**
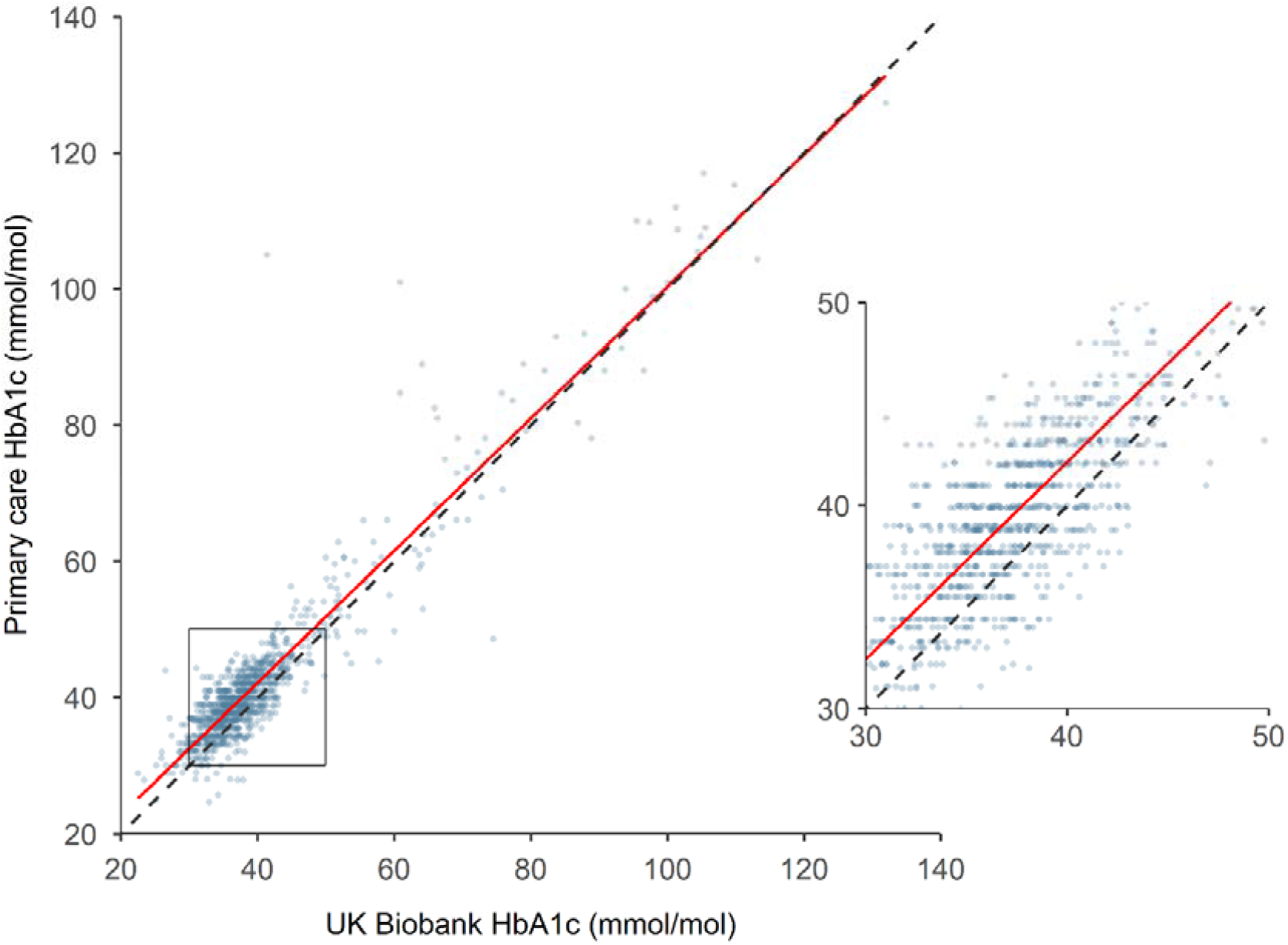
Scatter plot of HbA1c measurements taken at UK Biobank baseline assessment vs those in primary care taken ≤ 100 days before or after baseline assessment for *n*=1,039 individuals with no diagnosis of diabetes. Inset shows 30-50 mmol/mol values in more detail. Solid line: linear regression (equation: y = 0.9696x + 3.3595); dashed line: line of equality.

The difference in measurements from the two sources may be due to a number of factors. These include biological variation within an individual due to the time difference between the primary care measurement and baseline assessment (−100 to +100 days). However, there was not a significant association between this time difference and the difference in the measurement values (Pearson correlation coefficient = −0.025, *P* = 0.4; see Supplementary Figure S2). The difference in HbA1c values is therefore likely to be due to methodological differences. We consider the most probable contributors to be the use of different HbA1c analysers, and differences in sample storage (UK Biobank stored blood samples frozen for 4-10 years prior to analysing^4^). A brief discussion of these methodological differences and their potential contributions can be found in Supplementary Table S2. It should be noted that the UK Biobank and all NHS laboratories are registered with external quality assurance (EQA) schemes which verify the performance of HbA1c assays^3^, indicating that measurements from both sources meet the appropriate standards to be used for clinical decision making.

Small differences in measurements from different sources, which may not be clinically significant at an individual patient level, can result in large differences when used to define disease cases in a large cohort. Using HbA1c thresholds to identify cases of pre-diabetes (HbA1c 42.0-47.9 mmol/mol) and type 2 diabetes (HbA1c ≥ 48.0 mmol/mol) in this dataset results in under-diagnosis of pre-diabetes and diabetes when using UK Biobank HbA1c measurements compared to primary care HbA1c measurements, due to the differences between these measurements (Figure 2).

**Figure 2.**
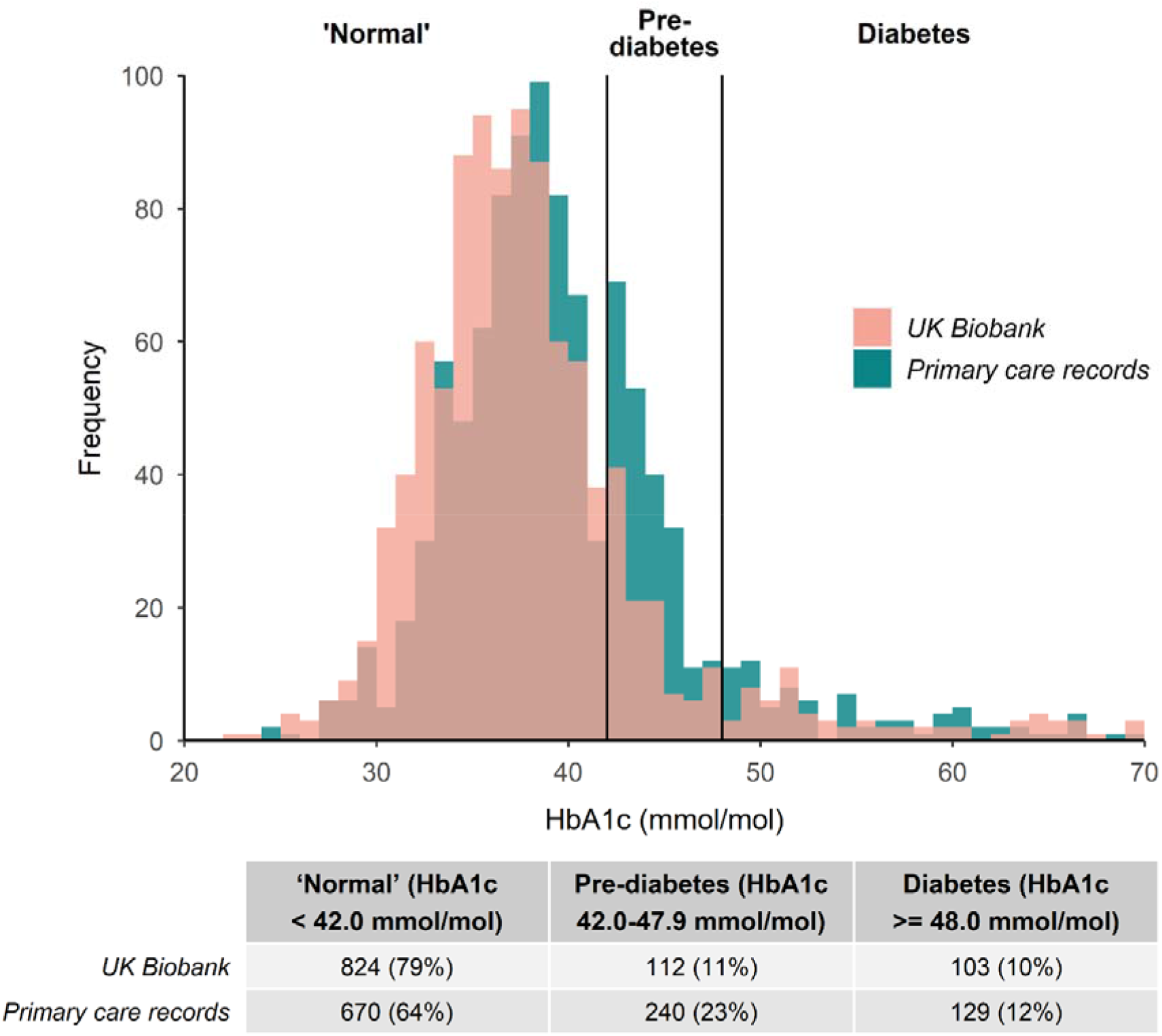
Histogram of HbA1c measurements taken at UK Biobank baseline assessment and in primary care records taken ≤ 100 days before or after baseline assessment for *n*=1,039 individuals with both measurements and no diagnosis of diabetes (x-axis truncated at 70 mmol/mol). HbA1c measurements in the ‘normal’ (< 42.0 mmol/mol), pre-diabetes (42.0-47.9 mmol/mol), and diabetes (≥ 48/0 mmol/mol) ranges are indicated; counts for these categories are shown underneath.

Participants who enter UK Biobank with a clinical diagnosis of type 2 diabetes will most likely have received this in primary care on the basis of an HbA1c measurement ≥ 48.0 mmol/mol. Additional cases of diabetes identified by a UK Biobank HbA1c ≥ 48.0 mmol/mol as in (5) cannot be treated as equivalent, because of the apparent difference between UK Biobank HbA1c measurements and primary care measurements. Re-aligning UK Biobank measurements using the equation of the linear regression line shown in Figure 1 as per Cull et al.^6^ may improve the comparability of these measurements. However, caution should be exercised as the *n*=1,039 primary care measurements in Figure 1 represent a range of different analysers, and only a subset of those used throughout the NHS for diagnostic diabetes testing.

Combining biochemical data from research studies with data from routine clinical care is becoming increasingly commonplace, whether for prospective cohorts such as UK Biobank or for randomised controlled trial participants^7^. Care must be taken to ensure that such data are equivalent and that it is valid to combine them. This is of particular importance when biomarkers are used to determine disease states, as in the case of HbA1c.

## Data Availability

UK Biobank data are available through a procedure described at http://www.ukbiobank.ac.uk/using-the-resource/.

## Ethics approval

UK Biobank received ethical approval from the North West Multi-centre Research Ethics Committee (REC reference: 11/NW/03820). All participants gave written informed consent.

## Author contributions

BMS and KGY conceived the study. KGY conducted the data analysis. All authors offered advice on the study design, analysis and interpretation of the results. KGY wrote the first draft of the manuscript. All authors read, reviewed, revised and approved the final manuscript.

## Supplementary data

Supplementary data are available at IJE online.

## Acknowledgments

This research has been conducted using the UK Biobank resource under Application Number 9072.

