## Supplementary for "Glycated haemoglobin measurements from UK Biobank are different to those in linked primary care records: implications for combining biochemistry data from research studies and routine clinical care"

### Supplementary Data

#### Supplementary Figure S1: Method for selecting UK Biobank participants with primary care HbA1c measurements within 100 days of baseline assessment

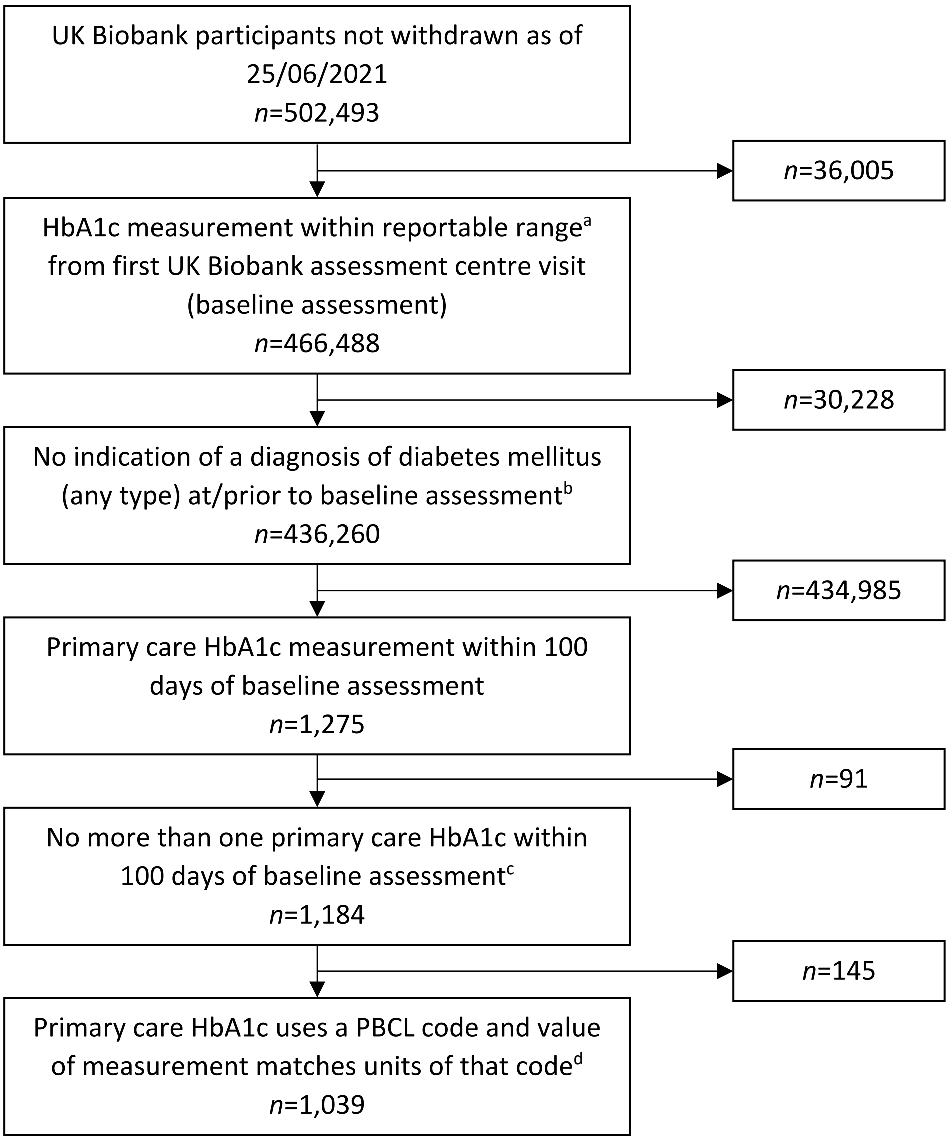


[a] Reportable range for UK Biobank HbA1c measurements was 15-184 mmol/mol (inclusive)[1].

[b] Participants with any indication of a diagnosis of diabetes mellitus (any type) at/prior to baseline assessment were removed in case they were receiving treatment resulting in rapid changes in HbA1c in the 100 day window between blood draws for the primary care and UK Biobank HbA1c measurement. Indications of a diabetes diagnosis were defined as:

- Diabetes diagnosis code in linked hospital episode statistics (HES) records with an episode start date prior to or on the same day as baseline assessment (admission date used where episode start missing)
- Diabetes code, HbA1c measurement ≥ 48 mmol/mol, or prescription for a glucose-lowering medication/glucagon/glucose testing strips in linked primary care records with a date prior to or on the same day as baseline assessment

- Self-reported diabetes, diabetes-specific complications, or prescription for glucose-lowering medications at first UK Biobank assessment centre visit (baseline assessment), or self-reported diabetes at second or third assessment centre visit where the diagnosis was before or in the same year as first assessment centre visit

Details of HES codes, primary care codes and UK Biobank self-reported variables used can be found in the Appendix of this document.

[c] Participants with more than one primary care HbA1c measurement within 100 days of baseline assessment were excluded as this close monitoring of HbA1c by their doctor may indicate that the participant was experiencing health issues affecting HbA1c or taking treatment expected to change their HbA1c. This did not include duplicate entries for a single HbA1c measurement (e.g. the same measurement recorded in both percentage and mmol/mol units) where these had the same event date.

[d] The Pathology Bounded Code List (PBCL) defines codes used in electronic reporting from pathology labs to general practitioners (GPs). Patient biomarker measurements with these codes may be more reliable than measurements with non-PBCL codes, as the latter are more likely to have been inputted manually by a GP. In order to include only reliable primary care HbA1c measurements in our analysis, we selected measurements with Read 2 and Read CTV3 PBCL codes specifically for HbA1c (not general 'glycosylated haemoglobin' codes or HbA1 codes) from the most recent PBCL release (October 2017; available via the NHS Digital TRUD website). The four codes selected are shown in Supplementary Table S1.

UK Biobank linked primary care data does not reliably include the units of biomarker measurements. We used the PBCL code description (Supplementary Table S1) to infer HbA1c measurement units (DCCT aligned = percentage units; IFCC aligned = mmol/mol units[2]), and then excluded measurements with a value outside of the clinically plausible limits for that unit[3].

Primary care HbA1c measurements in percentage were converted to mmol/mol using the NGSP/IFCC equation[2].

**Supplementary Figure S2: Scatter plot of time difference between blood draws for primary care HbA1c and UK Biobank HbA1c measurements vs difference in value of these HbA1c measurements (*n*=1,039)**

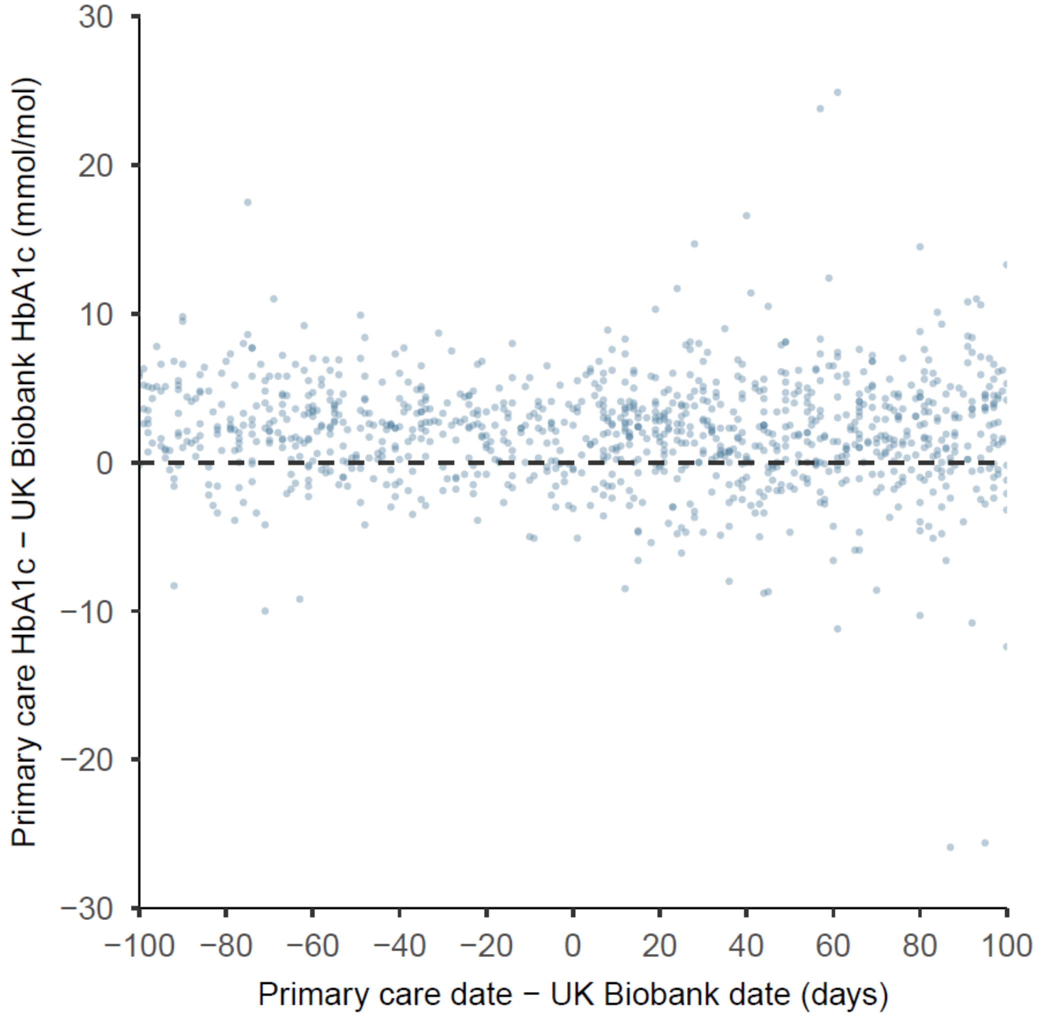


Pearson correlation coefficient = -0.025, *P* = 0.4

**Supplementary Table S1: Pathology Bounded Code List (PBCL) codes used to identify reliable primary care HbA1c measurements.** Measurements with values outside of the plausible limits were excluded.

| **PBCL code** | **Read 2 / Read CTV3** | **Code description** | **Unit[a]** | **Plausible limits of measurement value based on unit[3]** |
|---|---|---|---|---|
| 42W4. | Read 2 | HbA1c level (DCCT aligned) | percentage | ≥ 3.9 and ≤ 20 |
| 42W5. | Read 2 | HbA1c level (IFCC standardised) | mmol/mol | > 20 and ≤ 195 |
| XaERp | Read CTV3 | HbA1c level (DCCT aligned) | percentage | ≥ 3.9 and ≤ 20 |
| XaPbt | Read CTV3 | HbA1c level (IFCC standardised) | mmol/mol | > 20 and ≤ 195 |

[a] inferred from code descriptions: DCCT aligned = percentage units; IFCC aligned = mmol/mol units[2]

**Supplementary Table S2: Discussion of possible methodological differences between primary care and UK Biobank HbA1c measurements and their potential contributions to the differences observed in measurement values.**

Note: UK Biobank HbA1c measurements used venous blood samples. We expect the vast majority of HbA1c measurements in the primary care records to also be venous samples; although point-of-care testing (POCT) of capillary samples is available in some general practices, it is not recommended for diagnosis of diabetes[4]. Since our study population consisted of participants without a diagnosis of diabetes, we assume that the reason for their having an HbA1c test in primary care was screening for diabetes, and so POCT would not have been used. In addition, we have only used HbA1c measurements with Pathology Bounded Code List (PBCL) Read codes (Supplementary Figure S1), which are primarily from NHS laboratories, which only deal with HbA1c tests on venous samples.

| **Possible methodological difference** | **UK Biobank method** | **Primary care (NHS) method** | **Expected impact of difference** | **Contribution to observed differences[a]** |
|---|---|---|---|---|
| Storage of blood samples prior to HbA1c measurement | Whole blood sample in EDTA collection tubes stored at 4°C for < 36 hours and then in liquid nitrogen[5] for 4-10 years (median time for whole cohort of 482,331 with UK Biobank HbA1c measurement = 6.7 years)[b] | Whole blood sample in EDTA collection tubes stored at ambient temperatures for ≤ 72 hours and 4°C for ≤ 14 days[6] | Several studies have looked at the impact of freezing whole blood samples at -70 - -80°C on HbA1c measurement. Of the studies which stored samples for at least 12 months, both reductions[7-9] and increases[8] in HbA1c of 0.1-3.3 mmol/mol have been reported. The reason for these changes is unclear. | The small reductions in HbA1c value after storage at -70 to -80°C observed in previous studies[7-9] are comparable to the difference between UK Biobank and primary care measurements observed in this study (2.1 mmol/mol). Different storage methods could therefore feasibly account for this difference. |
| HbA1c analyser used | Five Bio-Rad Variant II Turbo HPLC analysers[1] | NHS laboratories use a range of analysers similar to that used by UK Biobank | External Quality Assurance (EQA) data from 2019-2020[10] show that Bio-Rad Variant II analysers, on average, obtain higher readings than the majority of analysers tested in the same EQA cycle, including the Arkray/Adams/ Menarini A1c HA-8000 and TOSOH HLC723/G7/G8/GX, which were the two most popular analysers. | Given the EQA results, we would expect the Bio-Rad Variant II Turbo used by UK Biobank to give higher readings than those seen in primary care, the opposite to what was observed in this study. We therefore do not think that analyser differences contributed to the observed differences. |

| | | | | |
|---|---|---|---|---|
| | | | Different analysers are affected differently by haemoglobin variants[11]. | The prevalence of haemoglobin variants in the UK population is relatively low (<5%[12]) and so is very unlikely to account for the differences observed in this study. |
| User differences (person performing HbA1c assays) | Insufficient information | Insufficient information | Could result in higher or lower readings | Insufficient information |
| Quality assurance/quality control protocols used | Registered with UK NEQAS EQA scheme[1]. ISO 17025:2005 Quality Accreditation[1]. | NHS laboratories are required to be registered with an EQA scheme such as NEQAS or RIQAS. | No difference expected | No difference expected |
| | Used algorithms to ensure that the order in which samples were tested was random in regards to geography, dates or time of day that the sample was collected[5]. This allowed identification and correction of day-to-day assay variation (correction was based on average assay values for each day)[5]. | Corrections for assay drift based on average assay result are not part of the routine quality control protocols used in NHS laboratories. | The date-of-assay corrections applied by UK Biobank were mostly very small (IQR -0.9 to +0.9 mmol/mol, median 0.0 mmol/mol). They had the overall effect of reducing higher values and increasing lower values, lowering the overall variability. | The UK Biobank date-of-assay corrections do not explain the difference between the UK Biobank and primary care HbA1c measurements. In the cohort used in this study (*n*=1,039), the raw (uncorrected) UK Biobank HbA1c measurements are on average 2.1 mmol/mol lower than the primary care HbA1c measurements (very similar to the 2.1 mmol/mol difference observed between the corrected values and the primary care measurements)[f]. |

[a] UK Biobank HbA1c measurements were lower by an average of 2.1 mmol/mol than those in primary care for a cohort for *n*=1,039 participants with a primary care HbA1c within 100 days of baseline assessment at UK Biobank.

[b] Uncorrected UK Biobank HbA1c measurements and dates of assays were from the Biomarker Assay Extended Dataset (Return 1602) provided by UK Biobank

**Appendix: Diabetes mellitus codes and variables**

**Hospital Episode Statistics (HES) and primary care codes**
All HES and primary care codes, together with notes on how they are implemented are available from the following public Github repository: https://github.com/drkgyoung/UK_Biobank_codelists.

**Self-reported UK Biobank fields**

| Field | Field description | Which assessment centre visit | Values used to indicate a diagnosis of diabetes prior to/at first assessment centre |
|---|---|---|---|
| 2443 | Diabetes diagnosed by a doctor | First | Any value other than 0 No (1 Yes, -1 Do not know, -3 Prefer not to answer, or missing) was treated as a potential indication of a diagnosis of diabetes |
| 6148 | Eye problems/ disorders | First | 1 Diabetes related eye disease |
| 6153 | Medication for cholesterol, blood pressure, diabetes, or take exogenous hormones | First | 3 Insulin |
| 6177 | Medication for cholesterol, blood pressure or diabetes | First | 3 Insulin |
| 20002 | Non-cancer illness code, self-reported | First | 1220 diabetes<br>1221 gestational diabetes<br>1222 type 1 diabetes<br>1223 type 2 diabetes<br>1276 diabetic eye disease<br>1468 diabetic neuropathy/ulcers<br>1607 diabetic nephropathy |
| 20002 | Non-cancer illness code, self-reported | Second and third | Any of the values shown above (1220/1221/1222/1223/1276/1468/1607) where year of diagnosis was prior to or the same as the year of the first assessment visit |
| 20003 | Treatment/ medication code | First | 1140857494 glibornuride<br>1140857496 glutril 25mg tablet<br>1140857500 glymidine<br>1140857502 gondafon 500mg tablet<br>1140857506 pramidex 500mg tablet<br>1140857584 acetohexamide<br>1140857586 dimelor 500mg tablet<br>1140857590 libanil 2.5mg tablet<br>1140874646 glipizide<br>1140874650 glibenese 5mg tablet<br>1140874652 minodiab 2.5mg tablet<br>1140874658 gliquidone<br>1140874660 glurenorm 30mg tablet<br>1140874664 tolazamide<br>1140874666 tolanase 100mg tablet<br>1140874674 tolbutamide<br>1140874678 glyconon 500mg tablet<br>1140874680 rastinon 500mg tablet<br>1140874686 glucophage 500mg tablet<br>1140874690 orabet 500mg tablet<br>1140874706 chlorpropamide<br>1140874712 diabinese 100mg tablet<br>1140874716 glymese 250mg tablet<br>1140874718 glibenclamide |

| | | | |
|---|---|---|---|
| | | | 1140874724 daonil 5mg tablet<br>1140874726 semi-daonil 2.5mg tablet<br>1140874728 euglucon 2.5mg tablet<br>1140874732 malix 2.5mg tablet<br>1140874736 diabetamide 2.5mg tablet<br>1140874740 calabren 2.5mg tablet<br>1140874744 gliclazide<br>1140874746 diamicron 80mg tablet<br>1140883066 insulin product<br>1140884600 metformin<br>1140921964 glucamet 500 tablet<br>1141152590 glimepiride<br>1141153254 troglitazone<br>1141153262 romozin 200mg tablet<br>1141156984 amaryl 1mg tablet<br>1141157284 glipizide product<br>1141168660 repaglinide<br>1141168668 novonorm 0.5mg tablet<br>1141169504 diaglyk 80mg tablet<br>1141171508 vivazide 80mg tablet<br>1141171646 pioglitazone<br>1141171652 actos 15mg tablet<br>1141173786 starlix 60mg tablet<br>1141173882 nateglinide<br>1141177600 rosiglitazone<br>1141177606 avandia 4mg tablet<br>1141189090 rosiglitazone 1mg / metformin 500mg tablet<br>1141189094 avandamet 1mg / 500mg tablet |
